# Comparative evaluation of EUCAST RAST and QuickMIC for rapid susceptibility testing of carbapenem-resistant organisms directly from positive blood cultures

**DOI:** 10.64898/2026.01.23.26344696

**Authors:** Nicole Degel-Brossmann, Tom Kimkes, Lucas Reibenspies, Jiabin Huang, Harald Seifert, Paul Higgins, Martin Christner, Martin Aepfelbacher, Cecilia Johansson, Christer Malmberg, Holger Rohde, Benjamin Berinson

**Affiliations:** Institute for Medical Microbiology, Virology and Hygiene, University Medical Center Hamburg-Eppendorf, Hamburg, Germany; Gradientech AB, Uppsala, Sweden; Uppsala University, Department of Medical Sciences, Uppsala, Sweden; Institute for Medical Microbiology, Immunology and Hygiene, Faculty of Medicine and University Hospital Cologne, University of Cologne, Germany; Institute of Translational Research, Cologne Excellence Cluster on Cellular Stress Responses in Aging-Associated Diseases (CECAD), Faculty of Medicine, University of Cologne, Cologne, Germany; German Centre for Infection Research, partner site Bonn-Cologne, Germany

**Keywords:** Rapid AST, EUCAST RAST, QuickMIC, blood culture, diagnostics, sepsis, resistance, comparative performance evaluation

## Abstract

**Objective:** The rapid availability of phenotypic antimicrobial susceptibility results is crucial for the timely detection of multidrug-resistant Gram-negative organisms and for guiding optimized treatment strategies. Recently, novel methods have been introduced that enable direct antimicrobial susceptibility testing (AST) from positive blood cultures. However, their performance has not yet been systematically compared in head-to-head evaluations. This study aimed to assess the analytical performance of two rapid AST approaches—the agar diffusion–based EUCAST rapid AST (RAST) method and the automated QuickMIC system—using a challenging collection of highly resistant Gram-negative organisms.

**Methods:** A total of 101 Gram-negative bacteria (*Escherichia coli*, n = 24; *Klebsiella pneumoniae*, n = 22; *Acinetobacter baumannii*, n = 30; *Pseudomonas aeruginosa*, n = 25) were spiked into blood cultures and processed according to the respective AST workflows. Broth microdilution (BMD) was performed from pure cultures as the reference method. Time to result (TTR), categorical agreement (CA), and essential agreement (EA) with BMD were evaluated. Boruta analysis was applied to identify genetic determinants associated with AST errors.

**Results:** Overall TTR for QuickMIC was 3 h 44 min with a CA of 86.2%, an EA of 92.3 % for Enterobacteriaceae and 97.0 % for non-fermenters. Overall CA of RAST ranged from 90.7%-93.7% across reading time points. Overall, very major discrepancy rates were low (QuickMIC n=0.7%, RAST n=0.1%). Presence of NDM-5 and KPC was most frequently associated with errors for QuickMIC and EUCAST RAST, respectively.

**Conclusions:** Both rapid AST approaches yielded robust results in this diverse and highly resistant bacterial study population, directly from positive blood cultures, with a short turnaround time. These findings underscore the potential of rapid AST methods to facilitate timely optimization of antimicrobial therapy in bloodstream infections, even in the context of extensively drug-resistant pathogens.

**Importance:** Accurate antimicrobial susceptibility testing (AST) is essential for stewardship and effective therapy, especially as rising antimicrobial resistance increases the risk of empiric treatment failure. Traditional AST methods are limited by slow turnaround times, creating a need for rapid alternatives. This study evaluated the diagnostic accuracy of two rapid AST methods—EUCAST RAST and QuickMIC—using 101 genetically characterized, carbapenem-resistant Enterobacterales, *Pseudomonas aeruginosa*, and *Acinetobacter baumannii* tested directly from positive blood cultures. Broth microdilution served as the reference. Both rapid assays provided results within 3.5–6 hours and demonstrated high categorical and essential agreement with few very major discrepancies. Incorrect results were more common in isolates harboring NDM-5 and KPC carbapenemases. Overall, the findings support EUCAST RAST and QuickMIC as reliable tools for challenging resistant pathogens and highlight their potential to enable earlier detection of carbapenem-resistant phenotypes and more timely initiation of appropriate, last-resort antimicrobial therapy.

## Introduction

The growing prevalence of infections caused by antimicrobial-resistant pathogens represents a major clinical challenge. In cases of severe infection and sepsis, antimicrobial resistance is of particular concern, as delays in the initiation of effective therapy are closely associated with rapid clinical deterioration and increased mortality[1–3]. Timely identification of the causative pathogen and its resistance profile is therefore of pivotal importance for the management of these patients. In recent years, several approaches for rapid species identification from positive blood cultures based on molecular as well as mass-spectrometry techniques have been developed[4, 5]. In clinically important Gram-positive pathogens, like *Staphylococcus aureus* or enterococci, identifying clearly defined resistance genes (*mecA/C* or *vanA/B*) causing resistance to methicillin or vancomycin, respectively, help to tailor appropriate therapy solely based on molecular results[6]. In Gram-negative pathogens, the substantial mechanistic and genetic heterogeneity underlying phenotypic resistance expression necessitates phenotypic antimicrobial susceptibility testing (AST), for which currently no alternatives exist[7].

Conventional AST is performed following overnight incubation of pure bacterial isolates, which results in prolonged turnaround times (TAT) and may contribute to extended periods of inappropriate antimicrobial therapy. In recent years, several rapid AST platforms (e.g., VITEK® Reveal™, Accelerate PhenoTest®, dRAST™) have been developed, providing results within approximately 5.5–7 hours. These methods shorten TAT and some have already demonstrated clinical utility in guiding timely and appropriate patient management[8, 9]. Nonetheless, these systems require dedicated instruments with potentially high acquisition costs, which may limit their implementation—particularly in low- and middle-income countries (LMICs), where the burden of AMR-associated mortality is greatest[7]. In this context, EUCAST RAST offers an attractive alternative, relying on a simple yet effective approach: spreading material from positive blood culture bottles onto agar plates with antibiotic disks and incubating them for predefined time points (4 h, 6 h, 8 h, and 16–20 h). Studies have demonstrated good concordance with broth microdilution (BMD) and reported a positive impact on patient management[10, 11]. However, the EUCAST protocol requires highly precise manual reading times and is currently limited to a small set of Gram-negative species, restricting its broader applicability. More recently, QuickMIC® (Gradientech AB, Sweden) was introduced to the market, combining a microfluidic and solid-phase cytometry approach. This technique covers a broader range of species than RAST resulting in automated phenotypic susceptibility results and interpretation (according to latest EUCAST or CLSI breakpoint tables) for 12 antibiotics in 2h-4h directly from a positive blood culture bottle[12, 13].

The development of several rapid AST approaches in recent years has resulted in a wide range of systems becoming available on the market. A direct comparison of these methods represents an urgent research need, particularly in the context of highly resistant bacterial populations, where patients are in critical need of rapid diagnostics to guide timely and appropriate treatment. Highly resistant isolates are especially challenging for AST methods, including rapid approaches. Therefore, the aim of this study was to evaluate the performance of EUCAST RAST and QuickMIC in comparison with reference BMD, using a contemporary collection of highly resistant isolates (carbapenem-resistant or carbapenemase-producing), which also underwent whole-genome sequencing. Specifically, we sought to assess overall performance and to determine whether certain carbapenem-resistance mechanisms pose particular challenges.

## Methods

### Characterization of the strain collection

A contemporary cryo-collection of 101 Gram-negative pathogens (*E. coli* n=24, *K. pneumoniae* n=22, *A. baumannii* n=30, *P. aeruginosa* n=25), carbapenem-resistant and/or carbapenemase-producing, was evaluated with EUCAST RAST and the QuickMIC system, and compared to BMD. Clinical isolates, collected between 2015 and 2024, which were tested as carbapenemase positive and/or carbapanem-resistant on standard of care diagnostic (SOC), including automated susceptibility testing using VITEK2 (bioMérieux, Marcy-l’Étoile, France). Species were identified by MALDI ToF MS analysis (Bruker Daltonics, Bremen, Germany).

The strain collection underwent whole-genome sequencing. Acquired resistance genes (ARGs) were identified using ResFinder[14, 15]. Boruta feature selection was employed to identify ARGs associated with discrepant phenotypic test results[16].

### Spiked blood cultures for RAST and QuickMIC

Bacterial isolates were spiked into blood culture (BC) bottles according to EUCAST RAST Quality Control (QC) criteria (“The European Committee on Antimicrobial Susceptibility Testing. Quality control criteria for the implementation of the RAST method. Version 5.0, 2022. http://www.eucast.org). BC bottles were incubated in a Bactec FX (BD, Heidelberg, Germany) automated BC incubating system until flagged positive. Positive BC were retrieved and further processed. For the EUCAST RAST method, BC bottles were lanced, and 125µL ±25 µL of BC medium were evenly distributed on a Mueller-Hinton (MH) agar plate (Oxoid, Basingstoke, United Kingdom) using a sterile swab. The following antimicrobial plates were applied to the agar plates: meropenem (MER) (10 µg), cefotaxime (CTA) (5 µg), ceftazidime (CTZ) (10 µg) (BD, Heidelberg, Germany) and ceftazidime-avibactam (CTV) (10-4 µg) (MAST Group, United Kingdom). Plates were incubated at 36 ± 1°C in ambient air, and zone diameters were read at time points 4 h, 6 h, 8 h, and 16-20 h and interpreted according to the corresponding EUCAST RAST breakpoints (version 6.1).

For automated susceptibility testing using the QuickMIC system, BC bottles were processed according to the manufacturer’s instructions. In brief, 10 µL of the positive blood culture fluid was added to the sample preparation vial containing melted agarose. The solution was filtered to remove red blood cells and subsequently injected in a QuickMIC cassette together with MH-II broth. After cooling in the refrigerator for 15 minutes, the cassette was placed in the QuickMIC device, where bacterial growth in the presence of various antibiotics was automatically assessed by the system, delivering MIC results after 3 h – 4 h. Results for the same four antibiotics evaluated with EUCAST RAST were used. MICs from QuickMIC were evaluated according to EUCAST AST breakpoints (version 15).

### Broth microdilution reference testing

Pre-filled BMD plates were used (Merlin Diagnostika GmbH, Bornheim-Hersel, Germany) and performed for all included isolates, following essentially ISO 20776-1:2019. The antibiotics, antibiotic abbreviations, and concentrations used are summarized in Supplementary Table 1. In brief, a 0.5 McFarland inoculum suspension prepared from overnight sub-cultured colonies were added to the plates, resulting in a final concentration of ∼5×10^5^ CFU/mL per well. The plates were incubated overnight in 37°C. The MIC was read after ∼20 h and interpreted according to the EUCAST breakpoints (version 15). Duplicate testing was used, and if the results did not agree a third replicate was tested and the modal result was reported.

### Data analysis

QuickMIC AST results, reference BMD MIC data, and EUCAST RAST category data were entered into the same database. ARG data were added by importing ResFinder export files and collating the identified genes and their overall classes. Essential agreement (QuickMIC–BMD) was calculated according to ISO20776-2:2021. Categorical agreement (CA), as well as minor, major, and very major discrepancies, were calculated as described previously[13]. The association between CA (for both rapid AST approaches vs. BMD) and ARG type was examined using Boruta analysis, where each ARG family was an independent variable and CA the dependent variable[16]. Boruta feature importance scores variables by repeated random forest analysis of all variables and their randomized “shadow” copies.

Variables with significantly higher importance than the best “shadow” are considered hits. Follow-up analysis assessed the direction of association. Genomes were annotated with PROKKA v1.14.5[17]. A pangenome was built with Roary v3.13.0 using default settings (95% protein identity) to derive a gene presence/absence matrix[18]. Core genes—defined as present in 99–100% of isolates by Roary—were individually aligned with MAFFT v7 and concatenated; the resulting multiple alignment was used to infer a maximum-likelihood phylogeny in RAxML-NG under GTR+G, with 1,000 bootstrap replicates and a random starting tree[19–21]. Trees were visualized with the R package ggtree in circular layout[22]; tip colors denote sequence type (ST), with MLST assigned via PubMLST/BIGSdb[23], for *A. baumanii*, using the Pasteur scheme; concentric heatmap rings indicate the presence (+) or absence (-) of resistance genes. All data were analysed in R (v4.4.2).

### Ethics

The collected bacterial isolates were not associated with any personal data from the source individuals, who were fully anonymous. No traceable or additional biological material was collected in this study. According to the Ethics Committee of the Hamburg Chamber of Physicians, no informed consent was required for the collection, analysis, and publication of these data.

## Results

### Genetic characterization of the dataset

100/101 bacterial isolates included for phenotypic susceptibility testing underwent whole-genome sequencing. Of note, NGS data from one isolate (*A. baumannii* - study number QRC005) could not be retrieved, however, PCR analysis demonstrated here the presence of an OXA-23 carbapenemase. Phylogenetic analysis employing cgMLST demonstrated presence of one clonal cluster of *A. baumannii* (comprising n=7 isolates) and three clonal clusters of *K. pneumoniae* (two clusters comprising two, one cluster comprising three isolates). Circular phylogenetic trees providing sequence types and per isolates absence/presence of antimicrobial resistance genes (ARGs) is provided in supplementary Figure S1.

Identification of ARGs by ResFinder, demonstrated the presence of a carbapenemase in 92/100 isolates that underwent WGS. All carbapenemase-negative isolates were identified as *P. aeruginosa* (QRC 11, 42, 46, 72, 74, 77, 94 and 118). Interestingly, these isolates all carried a frameshift mutation in *oprD* and additionally mutations in *mexR*, *nalC* and *nalD*, reasonably explaining the observed meropenem resistance. A histogram of all ARGs associated with carbapenem-non-susceptibility identified is depicted in Figure 1A. The association of ARGs and phenotypic resistance as determined by reference BMD is provided in Figure 1B. Interestingly, 14/24 *E. coli* isolates displayed non-resistant phenotype in BMD despite encoding for a carbapenemase (Oxa-48-like, n= 8; KPC n= 3; NDM, n= 2; VIM n= 1). In *K. pneumoniae* this was only the case for 4/22 isolates (Oxa-48-like n= 3, KPC n= 1).

**Figure 1:**
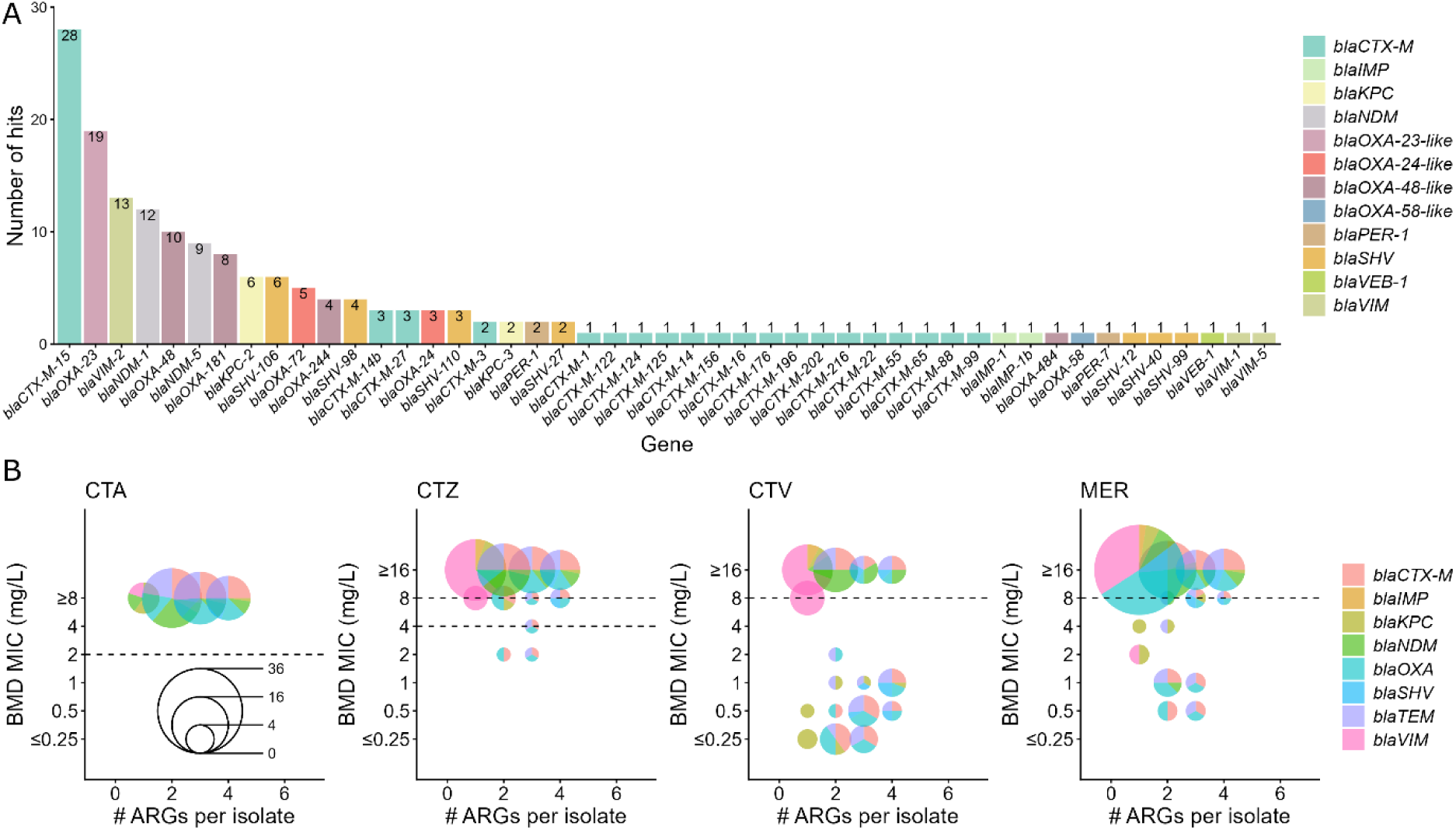
Histogram of ARGs and their effect on susceptibility results. **(A)** All antimicrobial resistance genes (ARGs) associated with carbapenem non-susceptibility, identified by ResFinder, are presented by gene and colour coded according to gene family. **(B)** Correlation between the presence of ARGs and reference broth microdilution (BMD) minimum inhibitory concentrations (MICs) for the four tested antibiotics. The size of the bubbles shows the number of isolates with the amount and type of ARGs encoded by colour in correlation to the corresponding MIC by BMD. CTA: cefotaxime; CTV: ceftazidime/avibactam; CTZ: ceftazidime; MER: meropenem. “R” breakpoint is indicated with a dotted line. In CTZ the lower line indicates the breakpoint for Enterobacterales and the upper line for *P. aeruginosa*.

### Analytical performance of RAST and QuickMIC

All 101 isolates were analysed using both RAST and QuickMIC. The average time to result (TTR) across all antibiotics for QuickMIC was 3 h 44 min. Specifically, the mean TTR for CTA, CTV, CTZ and MER were 3 h 50 min, 3 h 36 min, 3 h 48 min, and 3 h 46 min, respectively. Detailed raw data, including BMD results, QuickMIC results and TTR for each antibiotic, as well as RAST interpretations, are provided in Supplementary Table S2. CA results for CTA, CTV, CTZ, and MER were evaluated for Enterobacteriaceae (pooled *E. coli* and *K. pneumoniae*), *P. aeruginosa*, and the *A. baumannii*, which is shown in Figure 2. Overall CA of RAST compared to BMD at 4 h, 6 h, 8 h, and 16–20 h was 93.7 %, 91.4 %, 90.7 %, and 90.8 %, respectively, whereas overall CA for QuickMIC was 86.2 %. Both methods demonstrated high CA across most antibiotics. However, for meropenem, lower performance was observed for both RAST and QuickMIC. Specifically, overall CA for MER ranged from 77.1 % to 91.9 % across Enterobacteriaceae, *P. aeruginosa*, and *A. baumannii* for both systems. Notably, the QuickMIC system consistently produced a greater number of valid AST results compared to RAST. Early RAST readings were frequently invalidated due to difficulties in measuring zone diameters and the presence of the area of technical uncertainty (ATU); at 4h, QuickMIC generated 24.2 % more results than RAST. By 8h, this difference was largely resolved, with RAST producing only 1.1 % fewer results (Figure 2e). Overall, with RAST, zone diameters within the ATU accounted for 23.3 % of all non-interpretable measurements.

**Figure 2:**
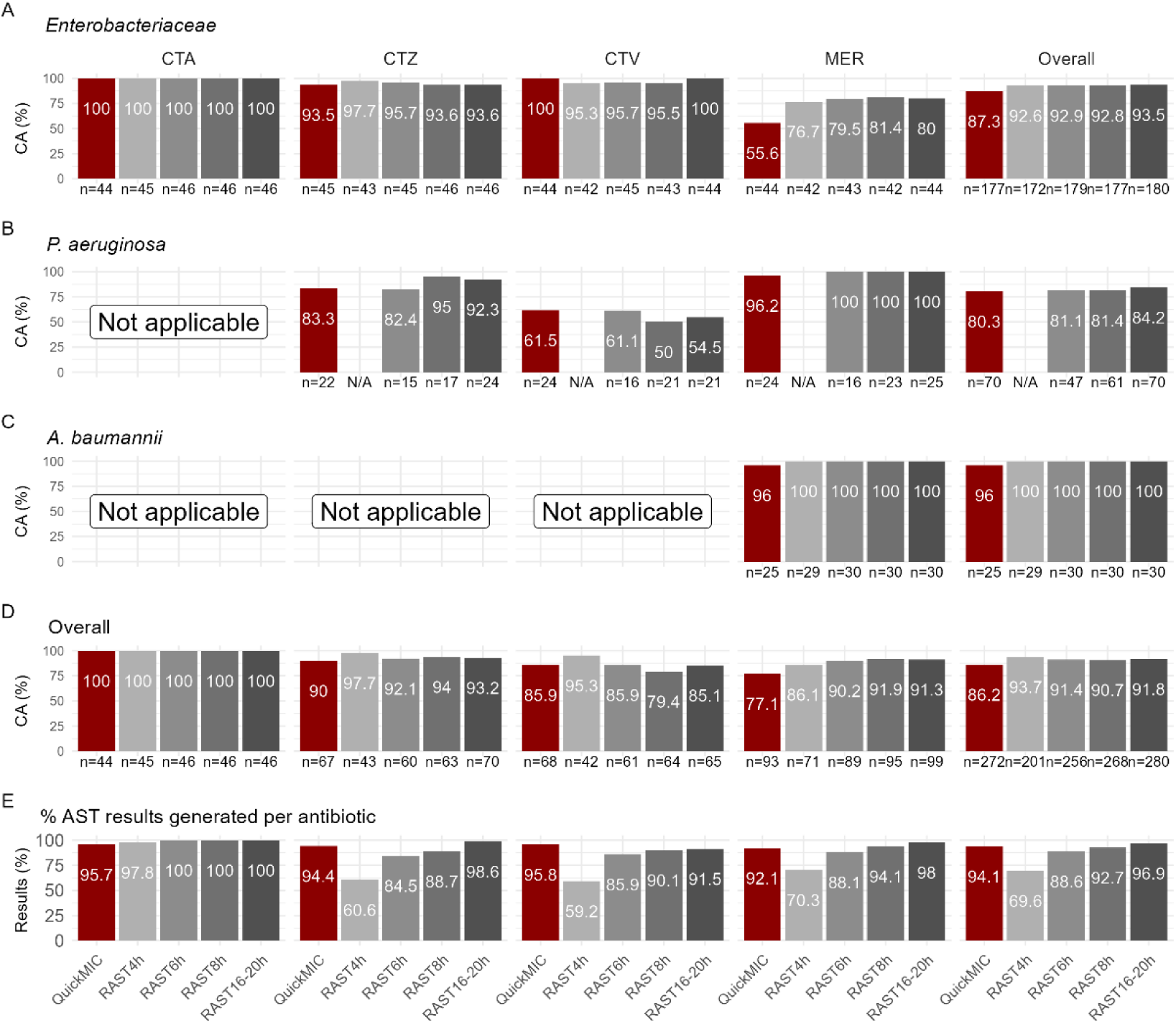
(A-D) Performance of QuickMIC and RAST per species using BMD as a reference. Bars indicate categorical agreement across all antibiotics tested (CTA: cefotaxime; CTV: ceftazidime/avibactam; CTZ: ceftazidime; MER: meropenem). QuickMIC results are indicated in red, RAST in grey, where shades of grey indicate results obtained for independent reading time points. **(A)** Enterobacteriaceae, **(B)** *P. aeruginosa* and **(C)** *A. baumannii* and **(D)** across all species. **(E)** Percentage of overall interpretable AST results available for each antibiotic and time point and system across all species. Colour codes correspond to A-D. N/A, not applicable.

CA discrepancies for both methods are summarized in Figure 3. Overall, discrepancy rates were low for both RAST and QuickMIC (VMD, MD and MiD for QuickMIC: 0.7%; 3.2% and 9.9%, respectively and for RAST: 0.1%, 4.4% and 3.7%, respectively). Of note, for MER and CTV the overall discrepancy rates were higher: for QuickMIC and meropenem: 1.0% VMD, 0.0% MD and 21.9% MiD; for RAST: 0.0% VMD, 3.3% MD and 6.5% MiD and for QuickMIC and CTV: VMD 1.4%, MD 12.7% and MiD 0.0%; for RAST: VMD 0.4%, MD 14.0% and MiD 0.0%. QuickMIC exhibited a higher minor discrepancy rate for Enterobacteriaceae with MER compared to RAST (42.2 % vs. 13.3–14.0 %). Performance for the other antibiotics and bacterial species was comparable between both systems. Of note, the majority (72.2%) of meropenem miD for Enterobacterales obtained with QuickMIC were attributable to isolates reported as “susceptible, increased exposure” (I) despite being categorized as resistant by BMD. Conversely, all miD observed with RAST resulted from isolates being classified as resistant, although they yielded an “I” category by BMD.

**Figure 3:**
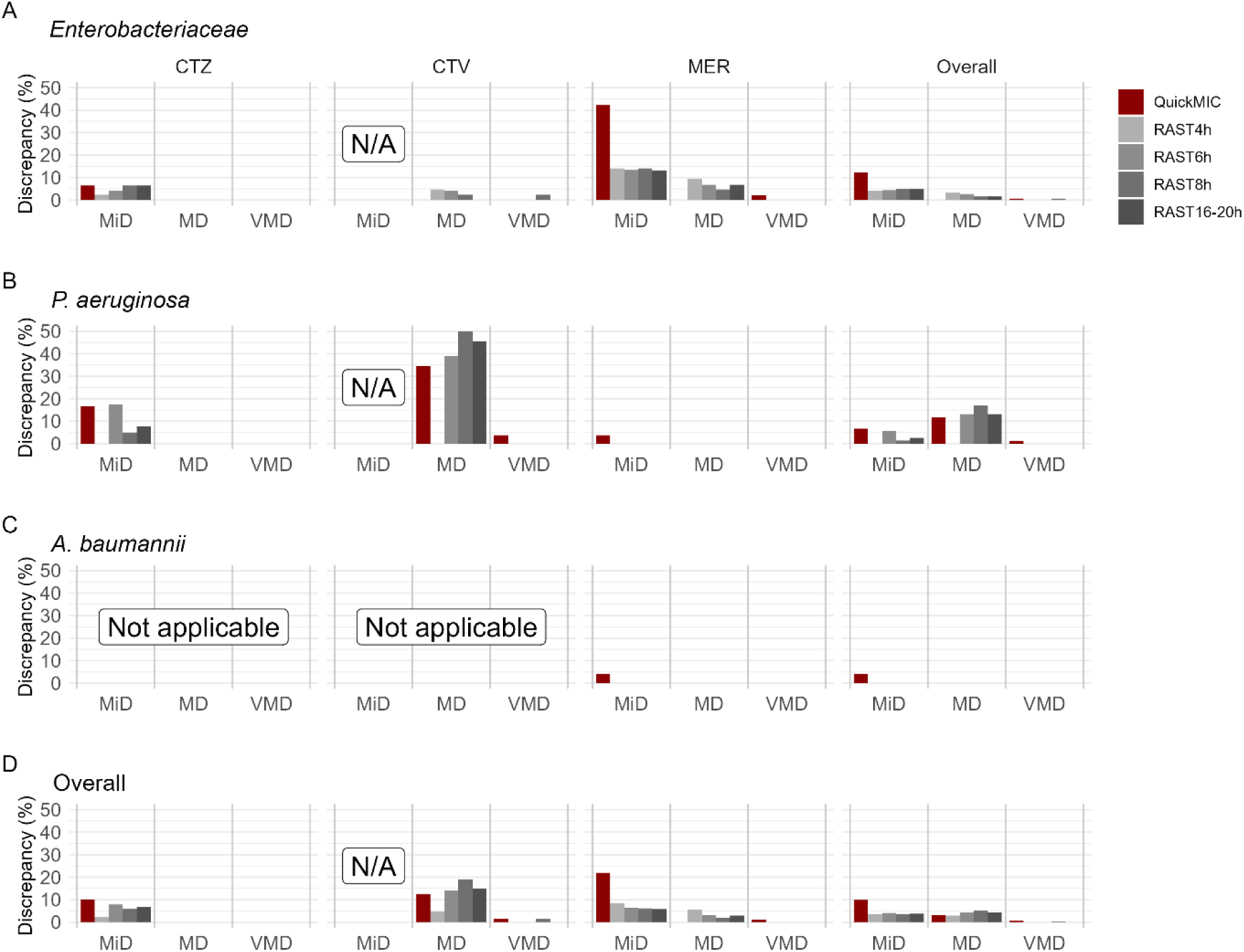
Overview of CA discrepancies per method and species. Bars indicate percentage discrepancies compared to BMD reference testing for the antibiotics tested (CTA: cefotaxime; CTV: ceftazidime/avibactam; CTZ: ceftazidime; MER: meropenem). QuickMIC results are indicated in red, RAST in grey, with shades indicating results at independent reading time points. MiD, Minor Discrepancy, MD, Major Discrepancy, VMD, Very major discrepancy. N/A, not applicable.

While RAST provides only categorical interpretations and therefore only CA can be evaluated, QuickMIC provides MIC values. MIC agreement with reference BMD is shown in Figure 4. Overall, essential agreement (EA) was 92.3 % for Enterobacteriaceae and 97.0 % for non-fermenters. EA exceeded 90 % for all tested antibiotics, with the exception of MER for Enterobacteriaceae (EA = 75.6 %).

**Figure 4:**
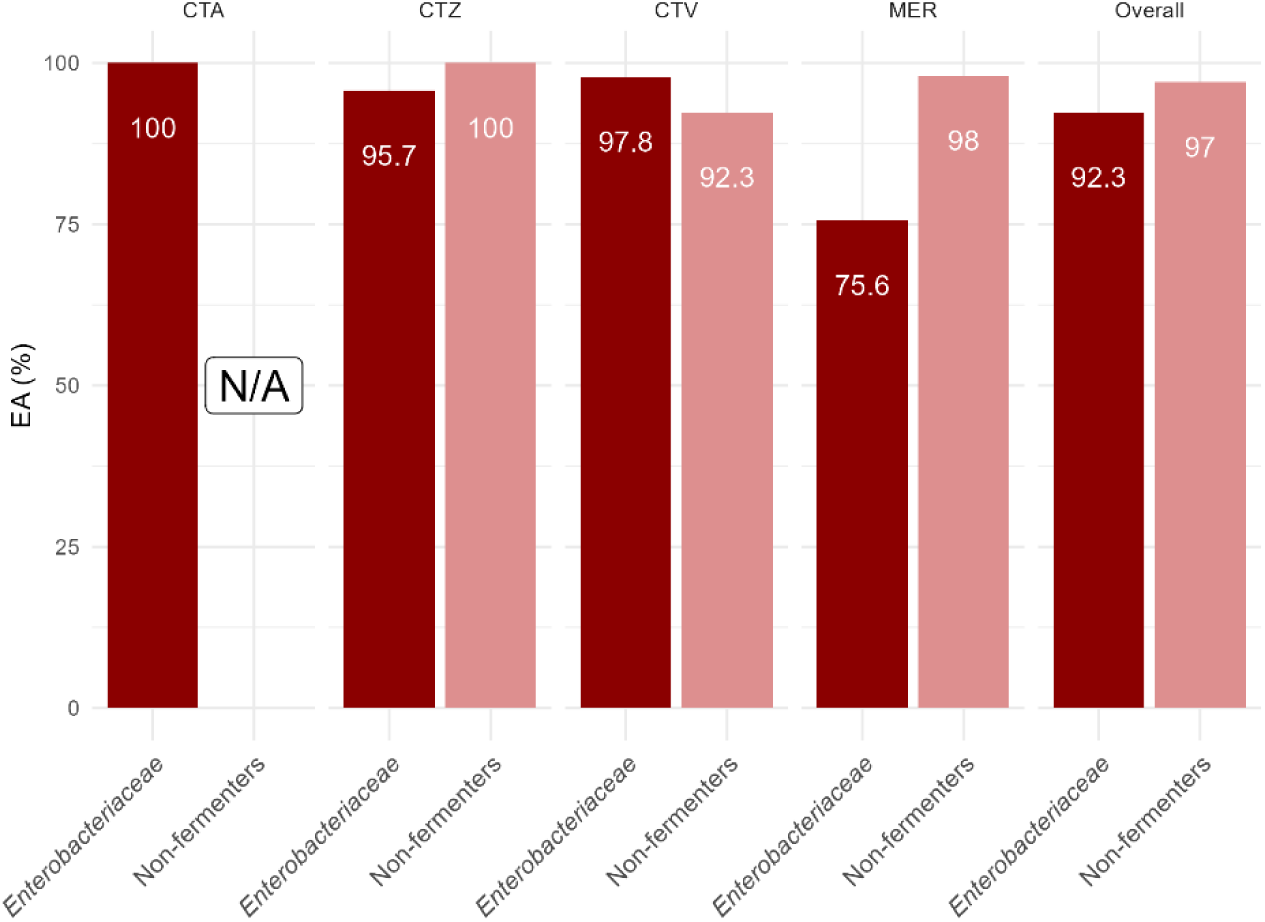
Overview of essential agreement for QuickMIC vs. BMD: EA rates for the four antibiotics investigated (CTA: cefotaxime; CTV: ceftazidime/avibactam; CTZ: ceftazidime; MER: meropenem), split up for Enterobacteriaceae and Non-fermenters in different shades of red for QuickMIC compared to BMD. N/A, not applicable.

### Genes associated with erroneous rapid AST results

Boruta analysis of ARGs and the association with type of ARG and CA error for MER is shown in Figure 5. In short, different ARGs were associated with performance limitations in the two methods. While presence of NDM-5 was most frequently linked to errors in QuickMIC, miss-classifications were associated with the detection of KPC for EUCAST RAST. A more detailed Boruta analysis for CA for QuickMIC and RAST at different time points for all tested antibiotics is presented in supplementary figure S2.

**Figure 5:**
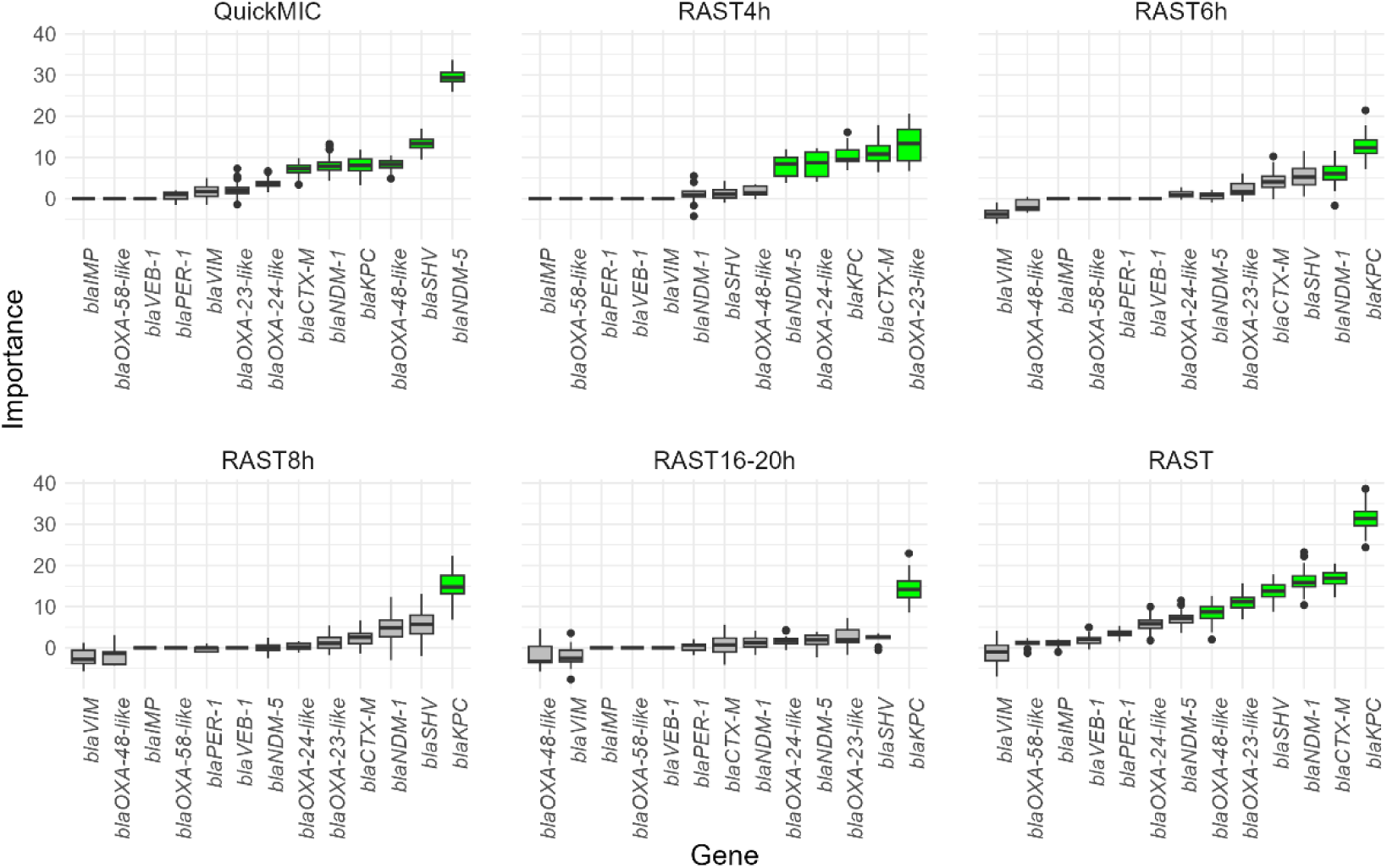
Boruta importance analysis for meropenem CA errors. Feature importance determined by Boruta analysis for the CA error with regard to meropenem for QuickMIC and RAST at different reading time points and overall. Each boxplot represents the distribution of importance scores for a specific resistance gene. Green boxes indicate features confirmed as statistically significant associations by the Boruta algorithm (p<0.01), while grey boxes represent non-significant features. The line in the boxes represents the mean importance.

## Discussion

An increasing number of rapid AST methods to allow susceptibility testing directly from positive blood cultures are becoming available. However, information on their specific strength and weaknesses is lacking, especially in the context of highly resistant Gram-negative rods. Therefore, here the performance of QuickMIC and EUCAST RAST was comparatively assessed using a collection of 101 carbapenem-non-susceptible and/or carbapenemase-producing Gram-negative isolates. Despite the high levels of resistance and the diversity of resistance mechanisms, compared to BMD both methods demonstrated robust performance, highlighted by CA of 93.7 % for RAST at 4 h and 86.2 % for QuickMIC. Both, QuickMIC and RAST reported low VMD rates (0.7%. and 0.1%, respectively). Interestingly, both methods performed better overall on non-fermenters than Enterobacteriaceae, a finding being in line with a recent study[24]. A key distinction between the two methods lies in turnaround time and result type. QuickMIC consistently generated MIC values with a mean TTR of less than 4 hours, providing a faster and more quantitative readout compared to RAST, which produced 24,2 % fewer interpretable results at the 4-hour time point and required longer incubation to achieve a complete dataset. This difference might have significant impact on the implementation of a rapid AST methods into routine work-flows.

Another important distinction between the two rapid AST methods is that EUCAST RAST provides defined screening cut-offs for the detection of ESBL- and carbapenemase-producing isolates, whereas QuickMIC does not offer such interpretive criteria (https://www.eucast.org/fileadmin/src/media/PDFs/EUCAST_files/RAST/2025/RAST_detection_of_resistance_mechanisms_v._3.0.pdf). This has important implications, as this might open opportunities to identify isolates carrying respective resistance genes, but are not resistant based on phenotypic AST. Indeed, 14 *E. coli* and 4 *K. pneumoniae* isolates, all carrying carbapenemase-genes, tested carbapenem non-resistant by BMD, but had MICs above the EUCAST screening cut-off. Analysis of the *E. coli* isolates revealed that all but one RAST measurement at the defined screening cut-off time points fell below the EUCAST-defined carbapenemase threshold (<20 mm zone diameter). Integrating these cut-offs with rapid gene detection methods—such as syndromic panel PCRs (e.g., BCID2, Biofire, bioMérieux, Marcy l’Etoile, France), in-house PCRs, or rapid phenotypic assays[4, 25]—could provide valuable information for the early identification of the underlying carbapenemase, potentially enabling timely optimization of therapy[26]. No screening-MICs are defined by EUCAST for rapid AST approaches and applying the general carbapenemase screening cut-offs (>0.125mg/L) is not feasible for the QuickMIC results, since the measuring range starts at 0.25mg/L. EUCAST currently recommends screening for carbapenemase-producers primarily for public health and infection control purposes, while otherwise reporting susceptibility results as observed (https://www.eucast.org/fileadmin/src/media/PDFs/EUCAST_files/Resistance_mechanisms/EUCAST_detection_of_resistance_mechanisms_170711.pdf). However, the interpretation of such results remains debated [27]. With regard to the already previously reported undercalling of the meropenem MICs [13], which was also noted here (72.2% of miD for meropenem in Enterobacterales, due to reporting “I” in QuickMIC, while being measured “R” in BMD), implies rigorous application of (molecular) testing methods, and careful interpretation of AST results. Optimal treatment strategies for patients infected with carbapenemase-producing organisms that appear susceptible to carbapenems remain unresolved and are the focus of ongoing research [28, 29].

A limitation of this study is that piperacillin-tazobactam (PIT) was excluded from this evaluation due to documented inherent problems of both methods to provide valid results[10, 11]. PIT is frequently used as first-line empirical therapy option in septic patients. Future studies should continue to address its inclusion and performance in rapid AST panels. Another limitation is the retrospective, monocentric study design, which may restrict the generalisability of findings to other clinical settings with different pathogen and resistance epidemiology. Moreover, no clinical data were collected, preventing assessment of the direct impact of rapid AST on treatment decisions, time to targeted therapy, or patient outcomes in infections with highly resistant isolates. The study also focused on a limited set of resistance mechanisms and cgMLST analysis indicated a small number of clonal clusters in *A. baumannii* and *K. pneumoniae*. Thus, broader validation across a larger strain collection, including additional resistance determinants, and diverse geographical regions is warranted. Finally, the use of whole-genome sequencing for resistance gene detection, while comprehensive, may not fully capture novel or uncharacterised mechanisms that could influence rapid AST performance.

The cost associated with implementation must be carefully considered. Direct cost associated for EUCAST RAST are reasonably low, making the method especially attractive for resource limited settings such as in LMICs. Integration of any automated method like the QuickMIC system is associated with high upfront cost related to the need to purchase a dedicated instrument, and higher cost per assay. However, the associated costs need to be weighed against specific and important advantages of automated systems, i.e. broader pathogen coverage and faster time to result. Moreover, fully automated reading and interpretation as well as integration into lab information systems reduce the manual workload, providing significant advantages related to lab work-flow integration and adherence to quality control standards. Importantly, the employment of rapid AST methods can lead to earlier targeted therapy and even improved patient outcomes[9, 11, 30]. Future studies need to assess if advantages of automated rapid AST methods will also improve patient outcome parameters, e.g., mortality, hospital length of stay, and reduction of broad-spectrum antibiotics usage. These might further make a business case for automated rapid AST methods.

### Transparency declaration

TK, LR, CM, CJ are Gradientech employees. HR is serving on the Gradientech scientific advisory board and received royalties for scientific presentations. BB received royalties from Gradientech for scientific advice. All other authors don’t declare any conflicts of interest.

## Authorship contribution

NDB acquired the data, performed analysis, writing of the draft.

TK and LR performed data analysis.

JH performed NGS analysis.

HS and PH provided species, draft editing and reviewing.

MC conceptualization, reviewing and editing of the manuscript.

MA reviewing and editing of the manuscript.

CM conceptualization, data analysis, writing, reviewing and editing of the manuscript.

CJ data acquisition

HR conceptualization of the study, writing, reviewing and editing of the manuscript.

BB conceptualization of the study, writing, reviewing and editing of the manuscript.

## Data availability

This Whole Genome Shotgun project has been deposited at DDBJ/ENA/GenBank under the genome accessions JBRZMQ000000000–JBRZQL000000000 (BioProject PRJNA1356847) and can be accessed via NCBI’s nuccore pages (https://www.ncbi.nlm.nih.gov/nuccore/JBRZMQ000000000) and the BioProject record (https://www.ncbi.nlm.nih.gov/bioproject/PRJNA1356847).

## Acknowledgements

The authors thank Rico Bluszis (UKE, Hamburg) and technical staff at Gradientech for excellent technical support.

---

**Figure S1.**
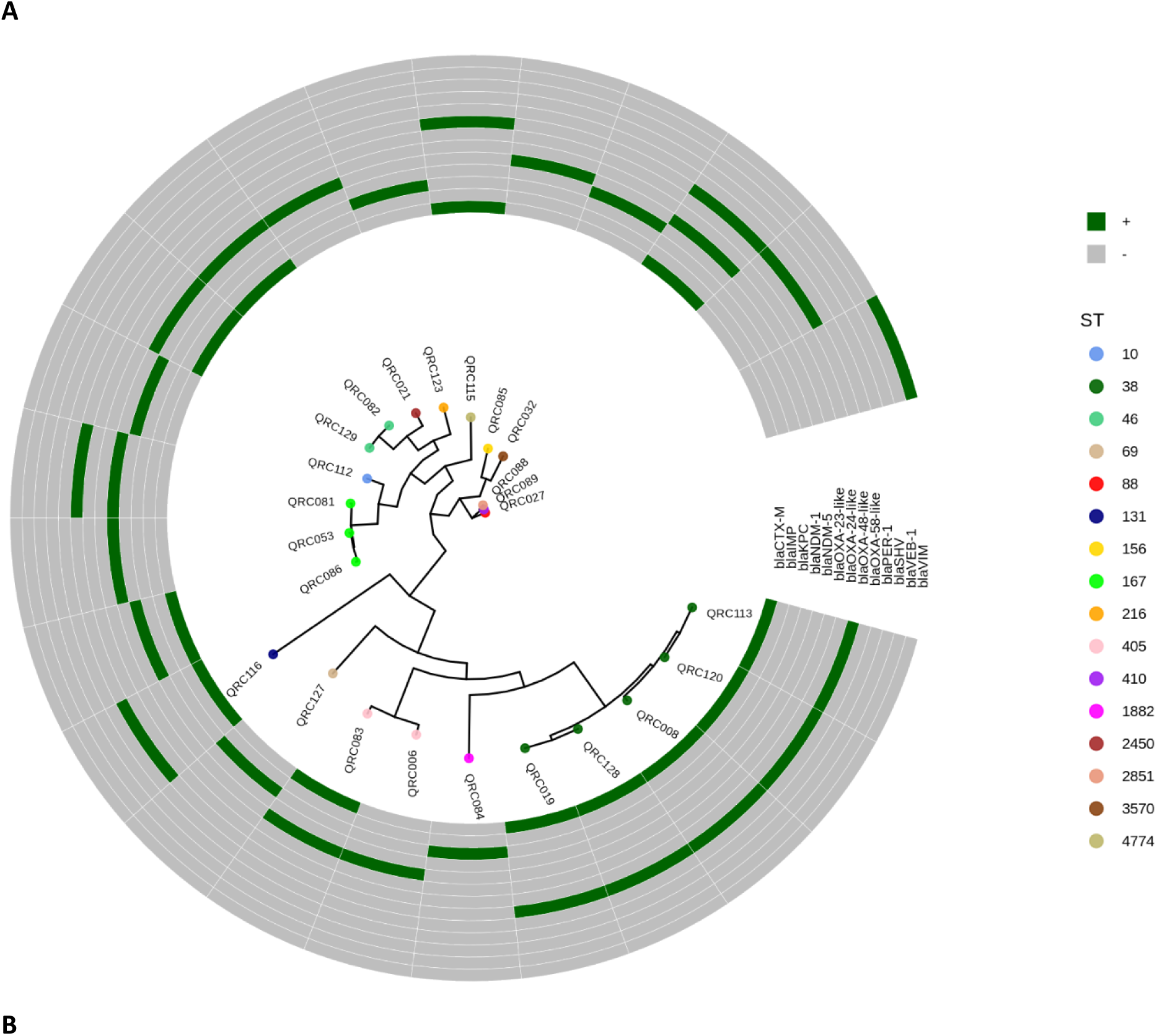
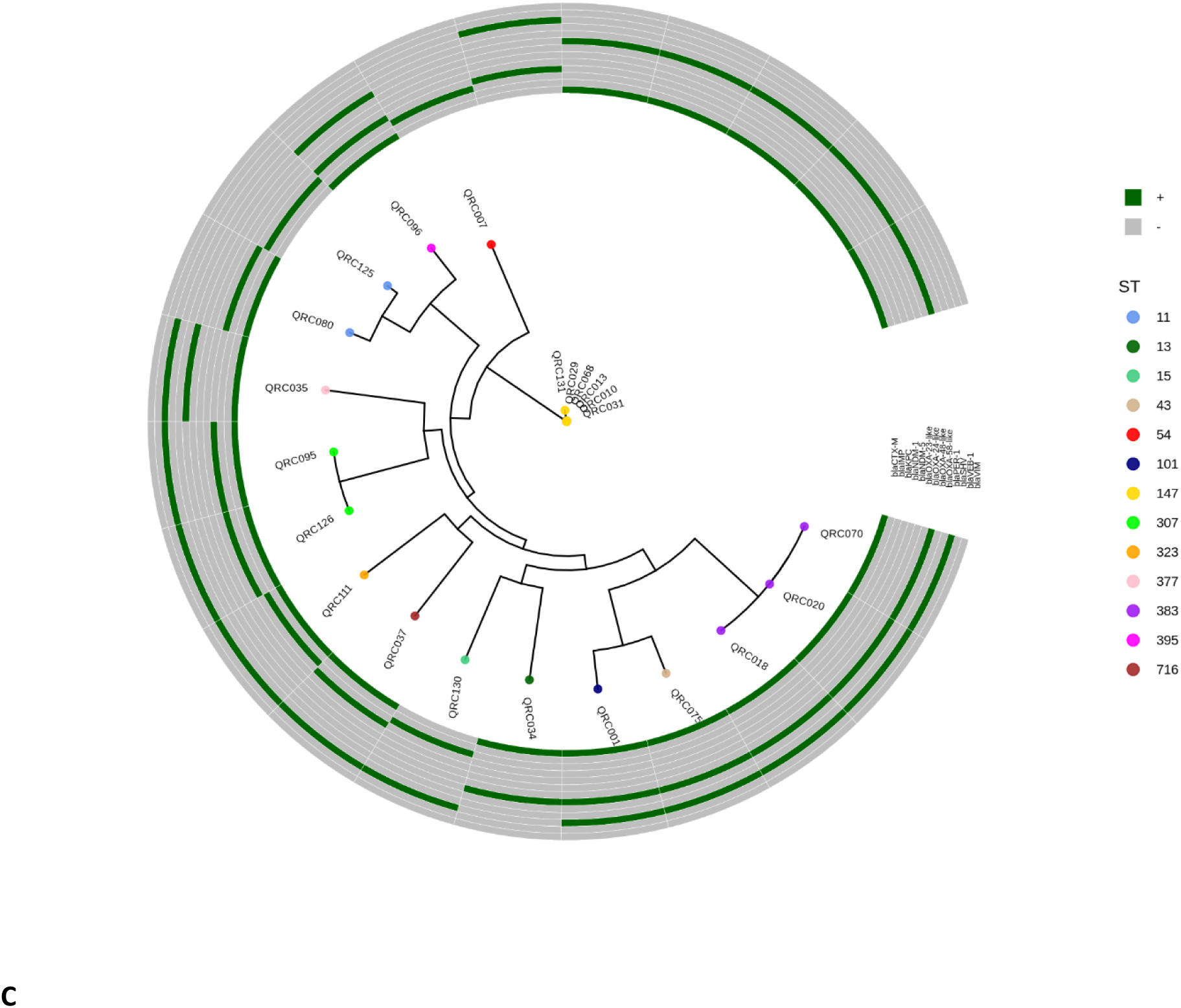
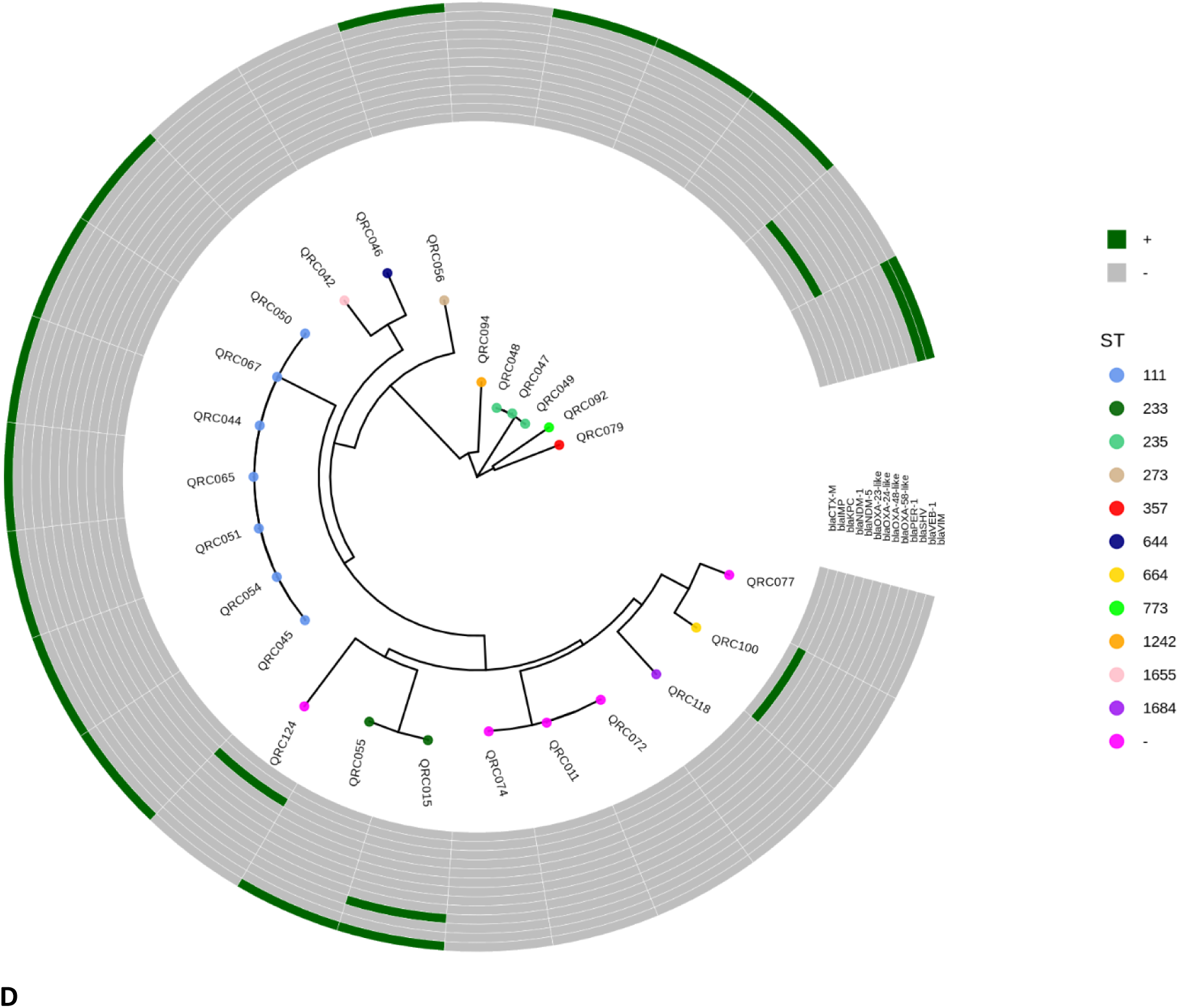
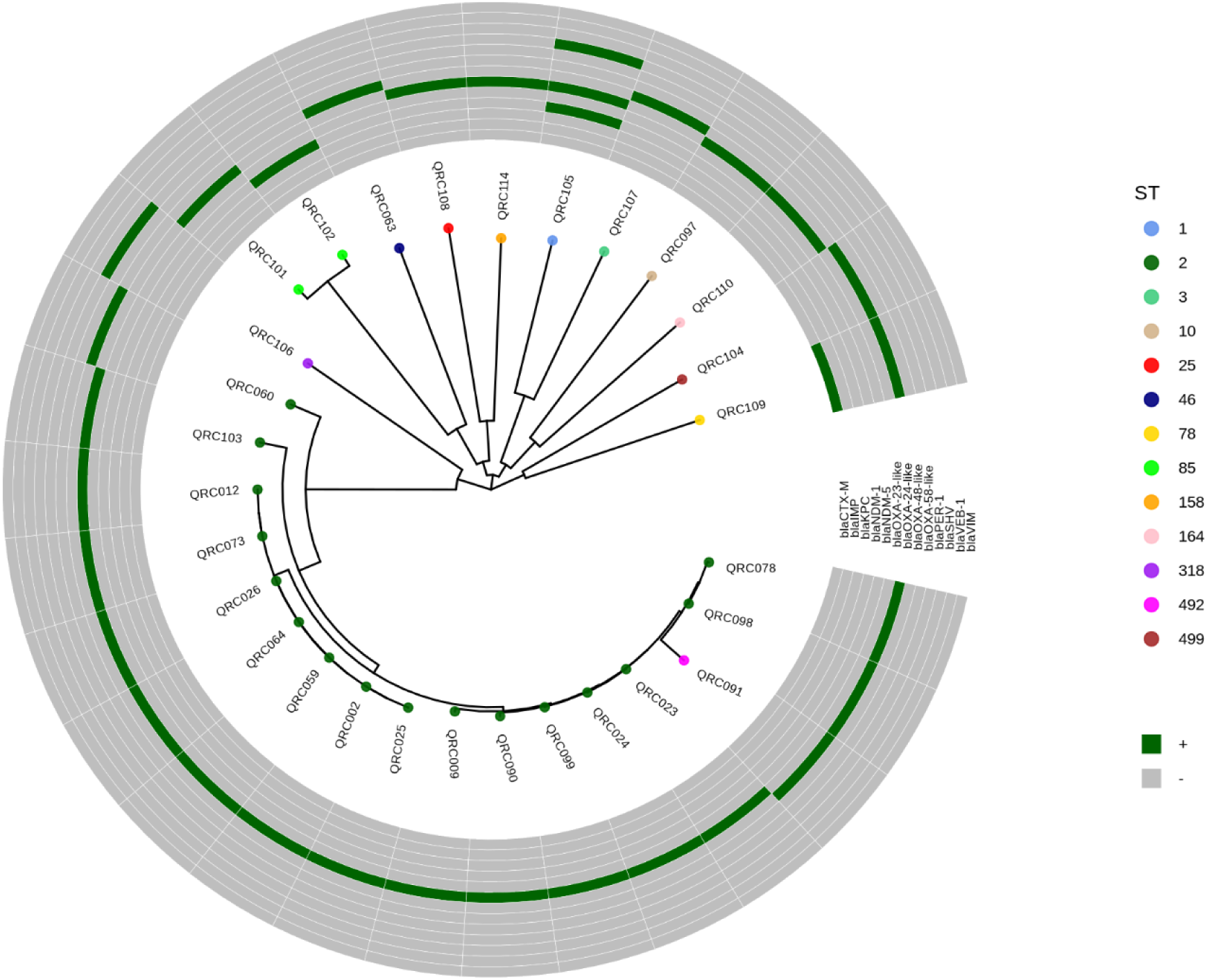
Phylogenetic analysis of study isolates using coregenome multi locus sequence typing (cgMLST). (A) *E. coli*; (B) *K. pneumoniae*; (C*) P. aeruginosa*; (D) *A. baumannii*. Using WGS, data a circular phylogenetic tree was constructed based on the cgMLST results. Sequence types are colour coded, according to the legend. Additionally, colour codes are indicating the presence (+; green) of absence (-; grey) of relevant ARGs

**Figure S2:**
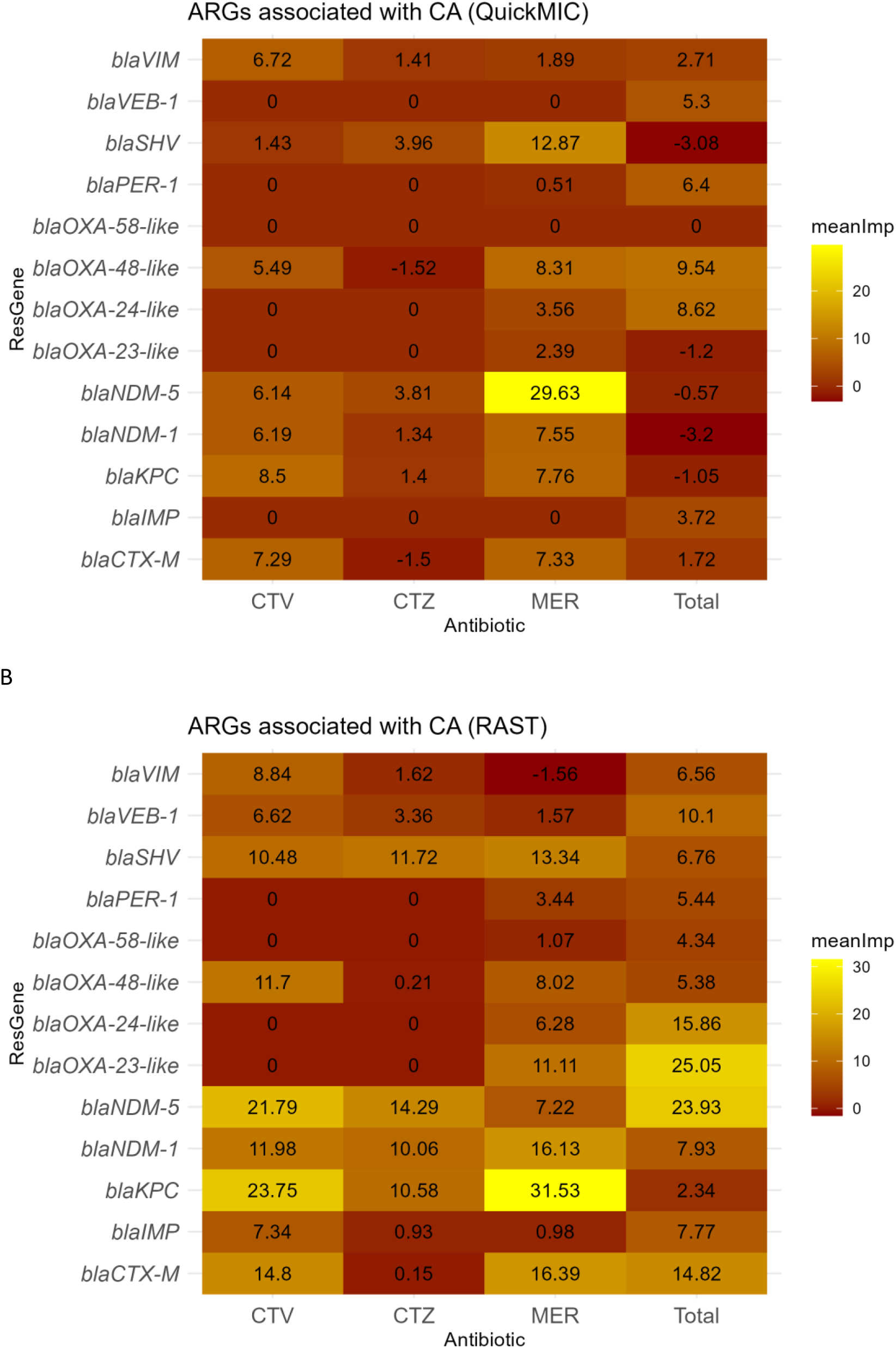
Heat map of Boruta analysis for different antibiotics. Boruta importance analysis of discrepancies of each gene cluster with regard to categorical agreement for (A) QuickMIC and (B) EUCAST RAST (all time points combined).

**Table S1.** Broth Microdilution plate measurement range and corresponding EUCAST breakpoints for Enterobacterales*, P. aeruginosa* and *A. baumanii*.

| Antibiotic |  | BMD<br>measuring<br>range (mg/L) | EUCAST<br>breakpoints<br>(v15)<br>(Enterobacterial<br>es, S/R) | EUCAST<br>breakpoints<br>(v15)<br>(*P. aeruginosa*,<br>S/R) | EUCAST<br>breakpoints<br>(v15)<br>(*Acinetobacter*<br>*species*, S/R) |
| --- | --- | --- | --- | --- | --- |
| **CTA** | Cefotaxime | 0.125-8 | 1/2 | - | - |
| **CTV** | Ceftazidime<br>/Avibactam | 0.25-16 | 8/8 | 8/8 | - |
| **CTZ** | Ceftazidime | 0.25-16 | 1/4 | 0.001/8 | - |
| **MER** | Meropenem | 0.25-16 | 2/8 | 2/8 | 2/8 |

**Table S2.**
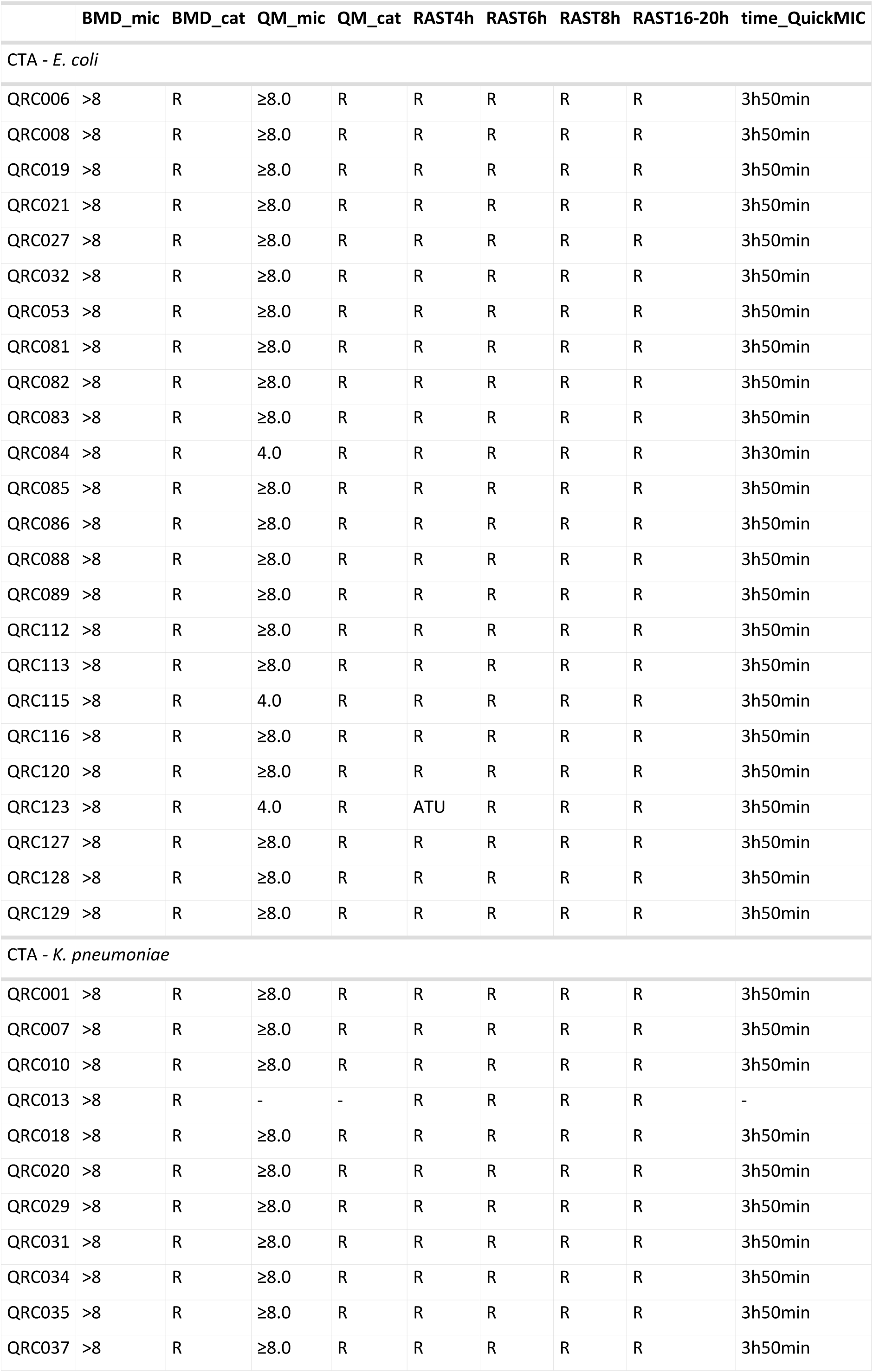
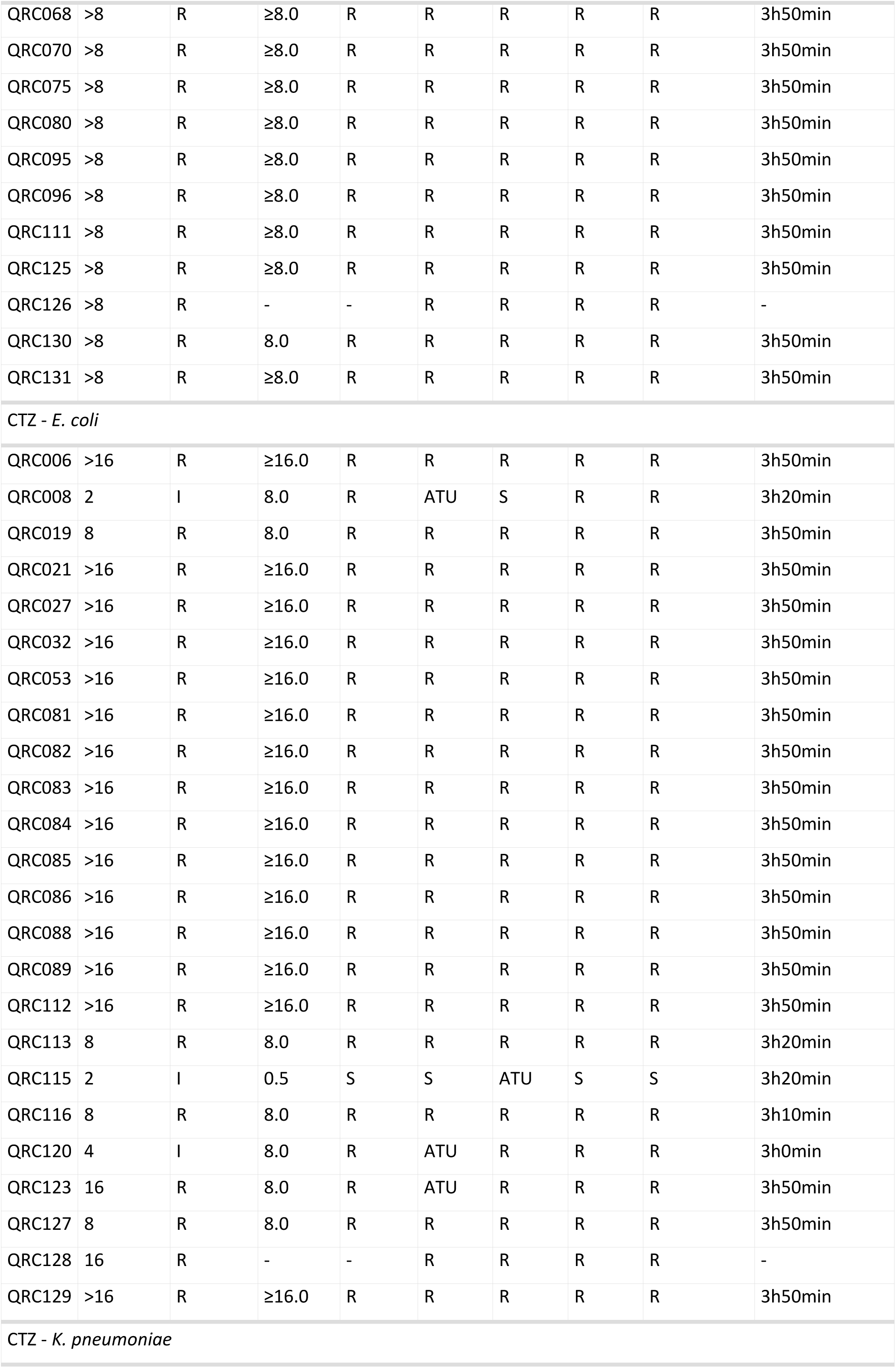
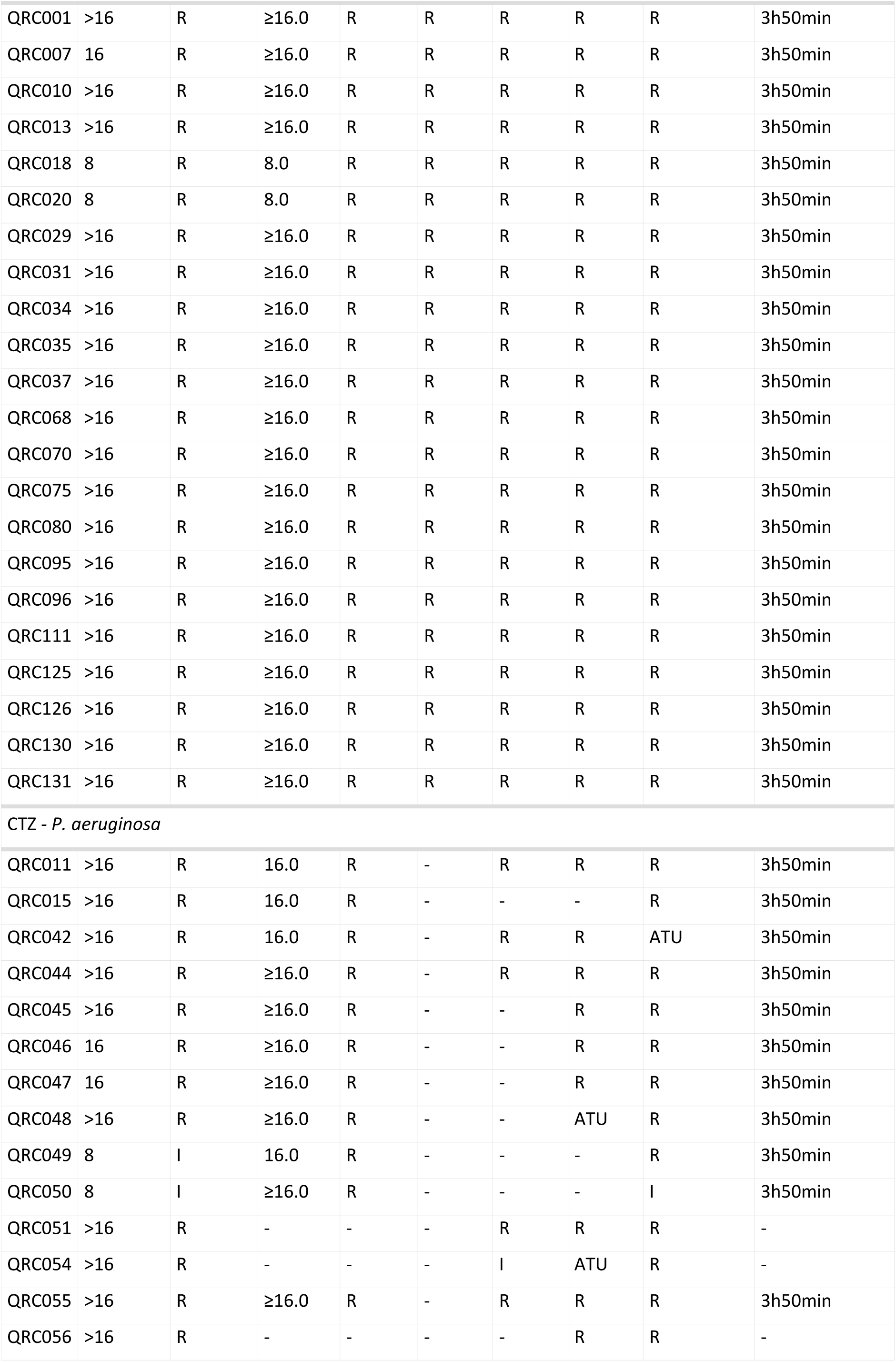
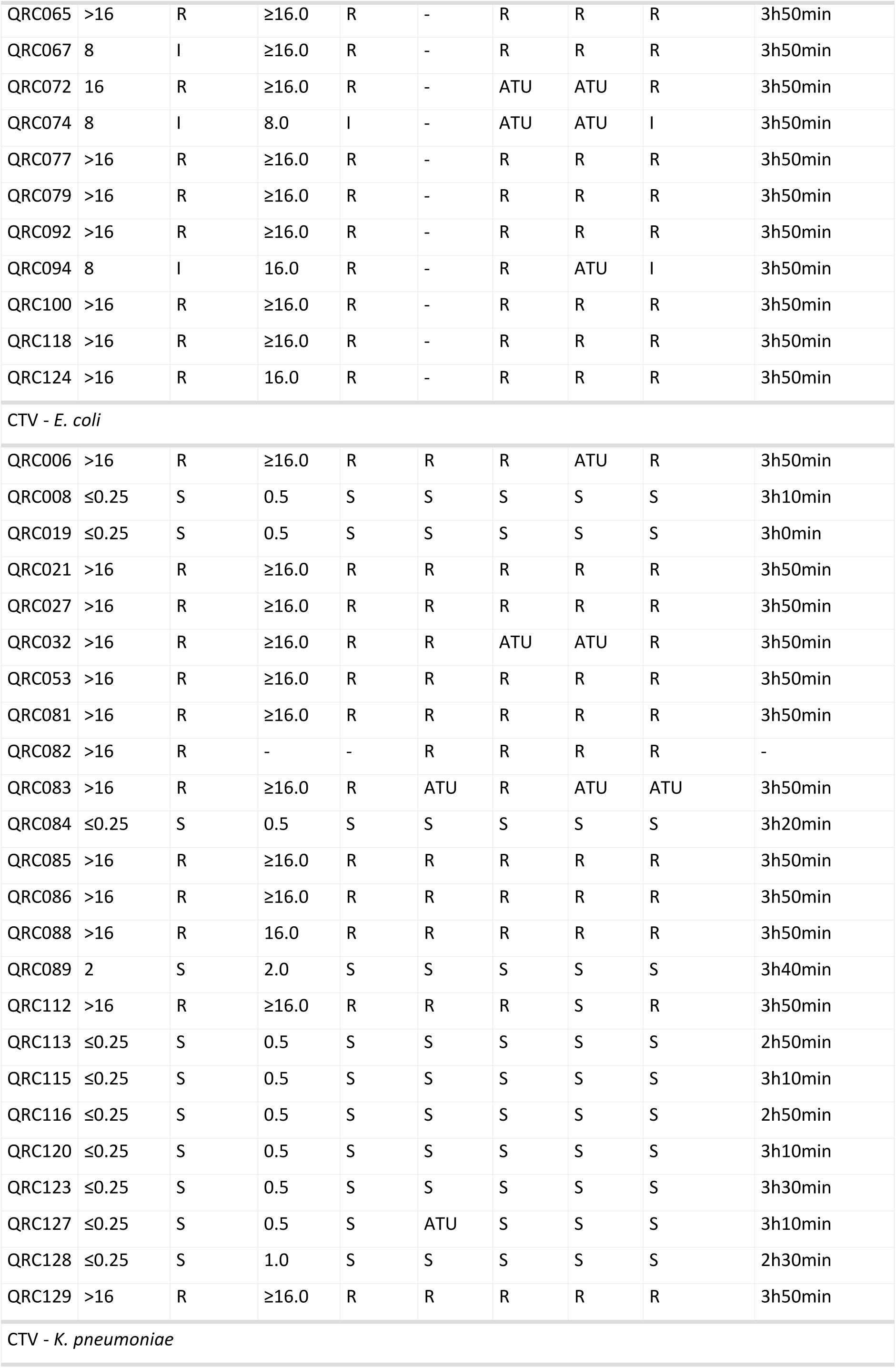
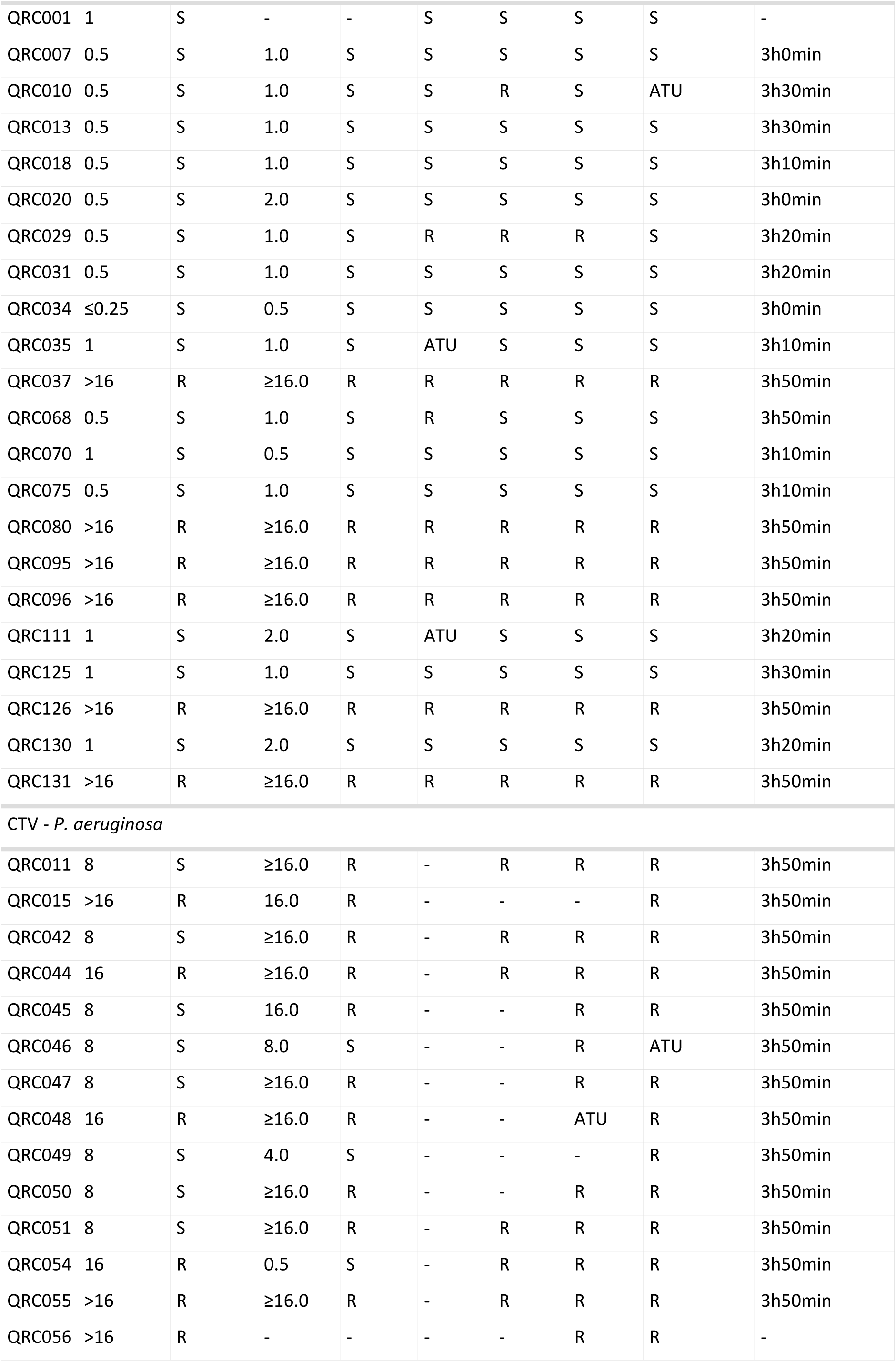
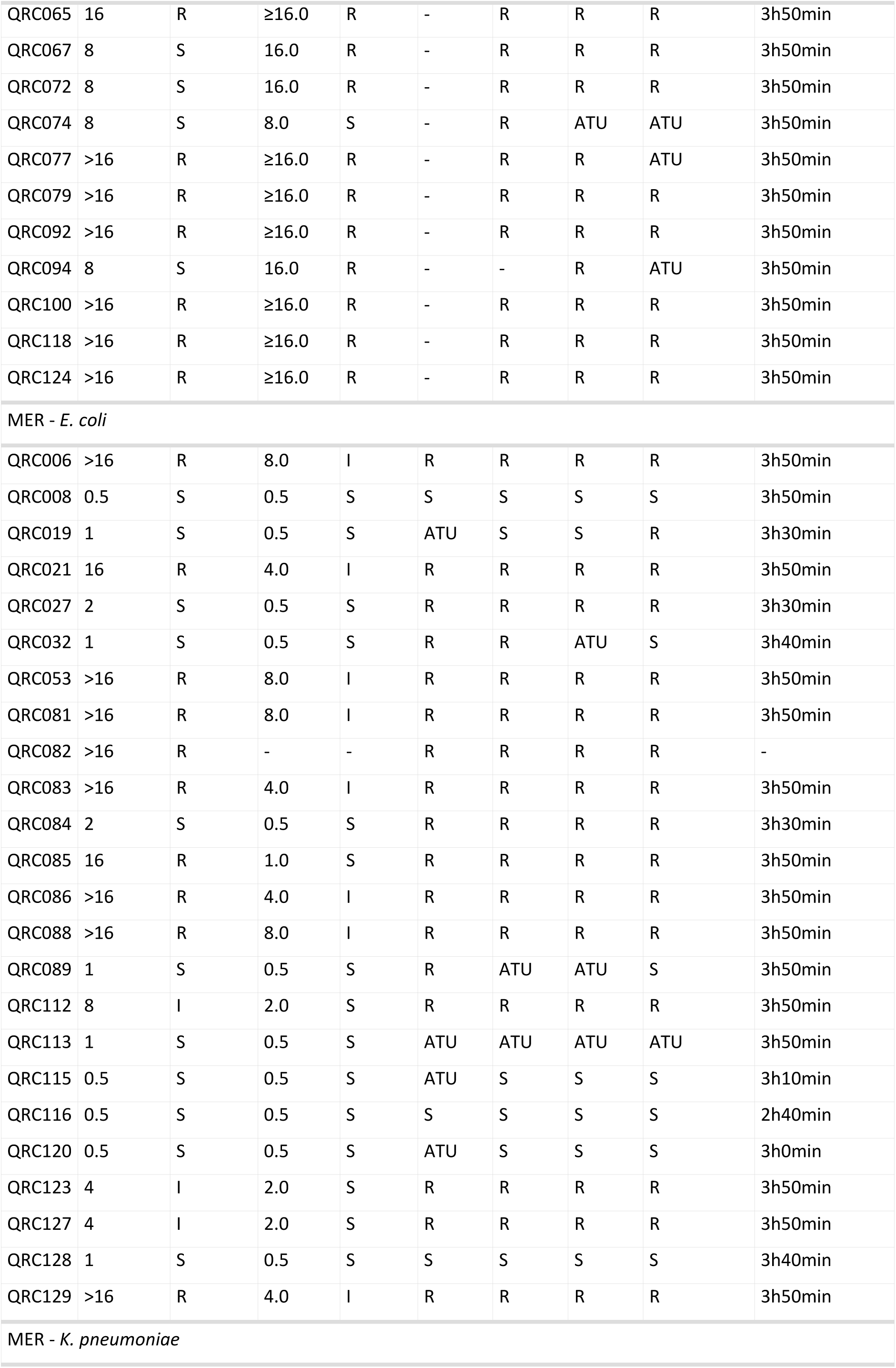
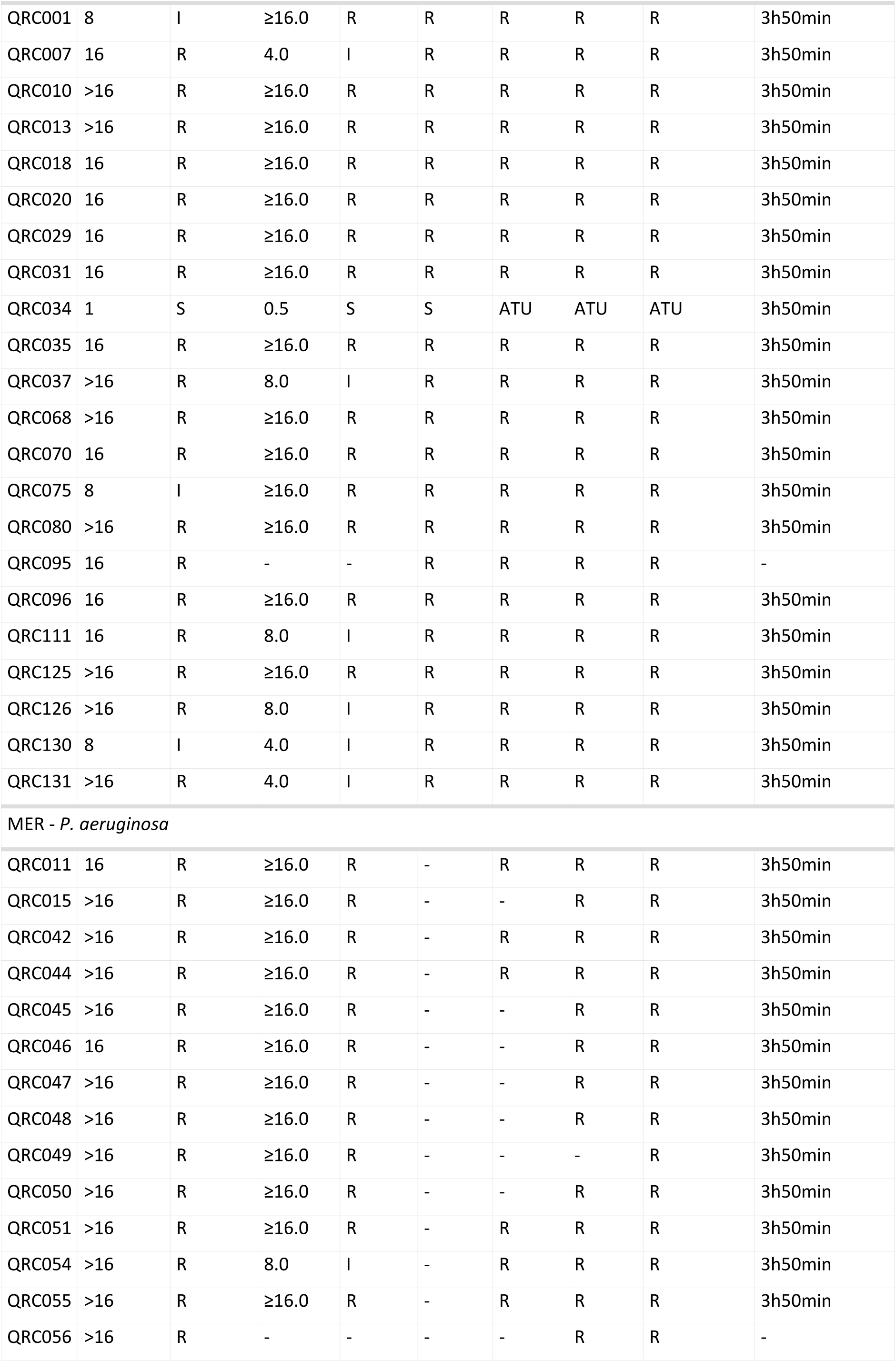
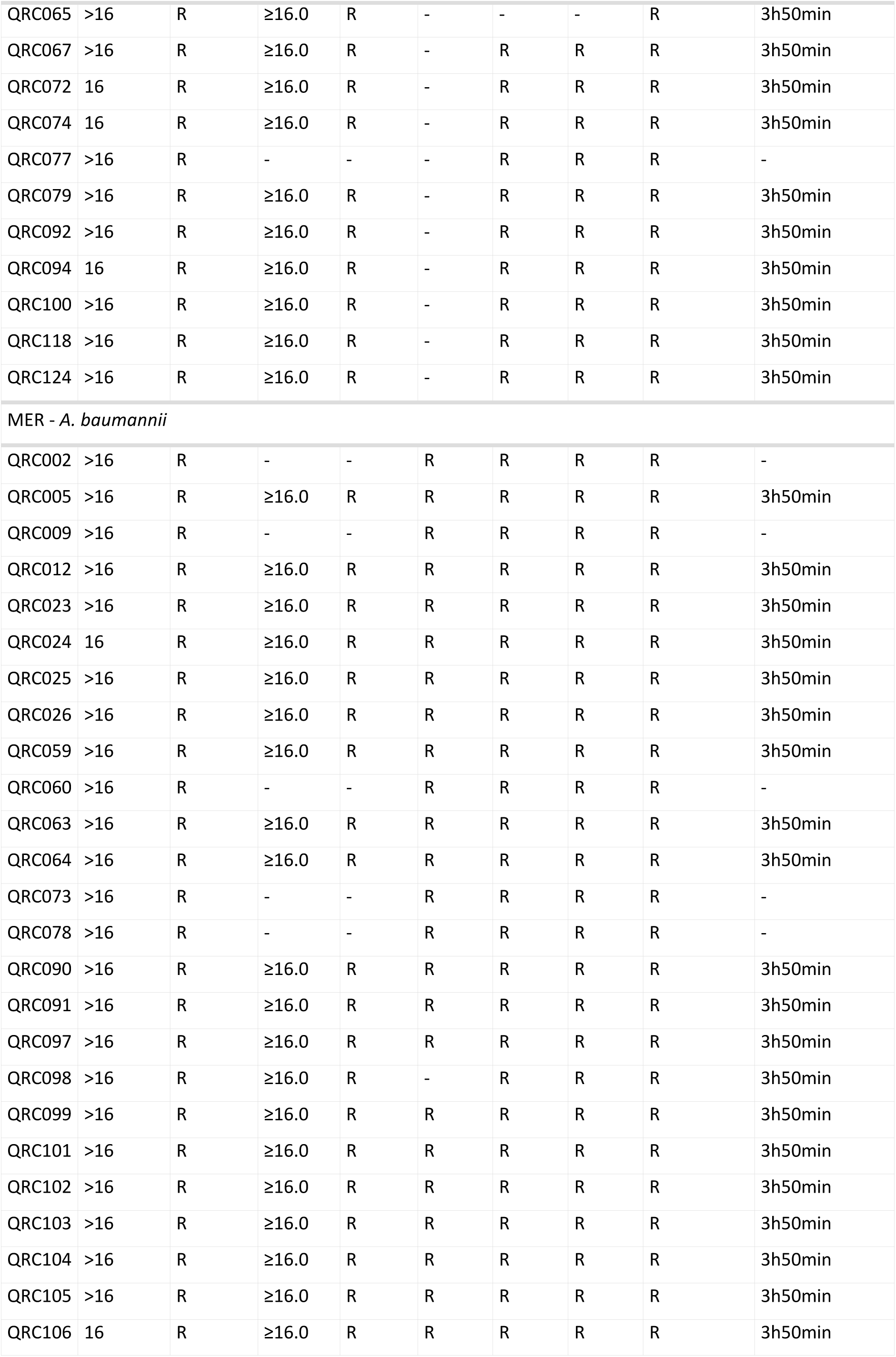
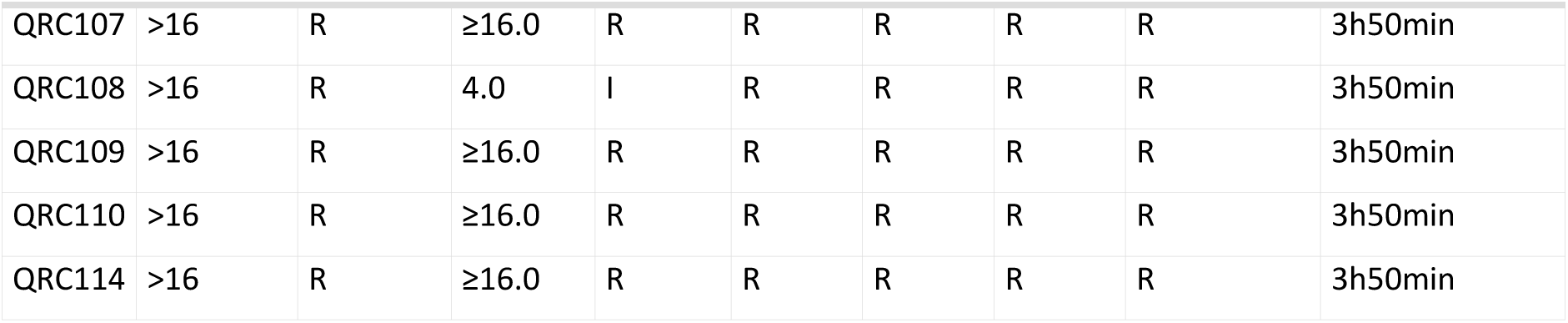
Overview of AST results for each strain.

